# Modeling COVID-19: Forecasting and analyzing the dynamics of the outbreak in Hubei and Turkey

**DOI:** 10.1101/2020.04.11.20061952

**Authors:** Ibrahim Halil Aslan, Mahir Demir, Michael Morgan Wise, Suzanne Lenhart

**Affiliations:** Department of Mathematics, Batman University, West Ramada, Batman, 72000, Turkey; Department of Fisheries and Wildlife/Quantitative Fishery Center, Michigan State University, East Lansing, 48823, MI, USA; Department Mathematics, University of Tennessee at Knoxville, Knoxville, 37996, TN, USA

**Keywords:** quarantine, social distancing, COVID-19 testing, novel coronavirus (COVID-19), reproductive number, forecasting, ordinary differential equations

## Abstract

As the pandemic of Coronavirus Disease 2019 (COVID-19) rages throughout the world, accurate modeling of the dynamics thereof is essential. However, since the availability and quality of data varies dramatically from region to region, accurate modeling directly from a global perspective is difficult, if not altogether impossible. Nevertheless, via local data collected by certain regions, it is possible to develop accurate local prediction tools, which may be coupled to develop global models.

In this study, we analyze the dynamics of local outbreaks of COVID-19 via a coupled system of ordinary differential equations (ODEs). Utilizing the large amount of data available from the ebbing outbreak in Hubei, China as a testbed, we estimate the basic reproductive number, *ℛ*_0_ of COVID-19 and predict the total cases, total deaths, and other features of the Hubei outbreak with a high level of accuracy. Through numerical experiments, we observe the effects of quarantine, social distancing, and COVID-19 testing on the dynamics of the outbreak. Using knowledge gleaned from the Hubei outbreak, we apply our model to analyze the dynamics of outbreak in Turkey. We provide forecasts for the peak of the outbreak and the total number of cases/deaths in Turkey, for varying levels of social distancing, quarantine, and COVID-19 testing.

## 1 Introduction

In late 2019, the city of Wuhan in the province of Hubei, China experienced an outbreak of Coronavirus Disease 2019 (COVID-19), the disease caused by the novel coronavirus SARS Coronavirus 2 (SARS-CoV-2). This outbreak quickly spread to all states of China and across the globe, being declared a pandemic by the World Health Organization (WHO) on 11 March 2020. The authorities imposed a strict lock-down on the city of Wuhan and other cities of the Hubei province on 23 January 2020 (World Health Organization, 2020b). In the face of over sixty-seven thousand cases and over three thousand deaths, the authorities continued strict enforcement of these measures (Chinese physicians, 2020; Coronavirus COVID-19 Global Cases by Johns Hopkins CSSE, 2020). Finally, on 23 March 2020, Hubei reached a significant milestone as the province’s health commission reported no new cases for seven consecutive days (World Health Organization, 2020b; The New York Times, 2020). Shortly thereafter, after over two months of severe restrictions on the movements of the Hubei population, the “2020 Hubei Lockdowns” were relaxed as the Hubei outbreak began to wane, inspiring hope that the global pandemic might be able to be controlled.

Previous studies of COVID-19 provided the evidence of human-to-human transmission and revealed its similarity and differences from SARS (Chan et al., 2020; Huang et al., 2020; Xu et al., 2020). However, data-driven simulation-based studies are needed to understand the dynamics of the ongoing outbreak. Indeed, it is of the utmost importance to use these tools to investigate the effectiveness of public health strategies, such as the number of COVID-19 tests carried out to detect the infected, the level of quarantine/social distancing, and its efficiency in the transmission of COVID-19. Many (preprint) studies investigate dynamics of this pandemic from a global perspective (see, e.g., (Imai, Dorigatti, Cori, Riley, & Ferguson, 2020; Read, Bridgen, Cummings, Ho, & Jewell, 2020; Riou & Althaus, 2020; Shen, Peng, Xiao, & Zhang, 2020; Zhao et al., 2020; Cao et al., 2020)). Nevertheless, the large variations in both quality and availability of data from region to region make direct global modeling of the dynamics of this pandemic exceedingly difficult.

As a result, in this study, we develop a model for dynamics of the pandemic from a local perspective. The many “hotspots” of COVID-19 combined with the many travel restrictions in place throughout the world further suggest that local models might provide more practical insights into the dynamics than their global counterparts. Indeed, it stands to reason that accurate models for local regions can be coupled to develop reasonable models for larger regions.

As of 23 March 2020, around one-quarter of the global COVID-19 cases and consequent deaths occurred in Hubei. The large proportion of data available from Hubei combined with the region’s recent achievements toward managing their local outbreak suggest that the data from this region presents an excellent picture of the lifetime of an outbreak of COVID-19. Indeed, as countries worldwide close their borders, cities and regions, and impose their own “shelter-in-place,” quarantine, or lockdown orders in the face of the pandemic, the large amount of data available from Hubei provides an excellent testbed for modeling the dynamics of a local outbreak of COVID-19.

In this study, we start by developing a SEIQR type deterministic model which uses a system of ordinary differential equations to analyze the dynamics of the outbreak, in particular highlighting the effect of testing and the effects of quarantine and social distancing in Hubei. We present estimates of the basic reproductive number *ℛ*_0_ of COVID-19 in Hubei and perform a sensitivity analysis to deduce which parameters play significant roles in the transmission and control of the outbreak in Hubei. In addition, we also provide 15-day forecasts of the fatality rate of the outbreak, the number of cases, and the number of deaths depending on the data (Chinese physicians, 2020; Coronavirus COVID-19 Global Cases by Johns Hopkins CSSE, 2020; World Health Organization, 2020b) and outputs of our SEIQR model. Finally, building on knowledge obtained from the Hubei outbreak, we apply our model to the outbreak in Turkey. We forecast the peak of the outbreak and the total number of cases/deaths in Turkey, utilizing the extant COVID-19 data from Turkey ((Ministry of Health (Turkey), 2020)).

## 2 Model formulation

A deterministic compartmental model has been developed by using ordinary differential equations (ODEs) to understand the dynamics of COVID-19 in Hubei, China (Chubb & Jacobsen, 2010; Keeling & Rohani, 2008; Kot, 2001). In the model, the total population *N* (*t*) at time *t* is divided into the following six compartments: susceptible *S*(*t*), susceptible in quarantine (isolated class) *S*_*q*_(*t*), exposed *E*(*t*), infected (asymptomatic or having mild symptoms) *I*(*t*), reported (infected) cases (hospitalized if get severe symptoms or quarantined if get mild symptoms) *I*_*q*_(*t*), and recovered *R*(*t*). Note that all individuals who, upon testing, test positive are immediately isolated. The transition flows among compartments are given in Figure 1. The rate of reported cases *i*_*q*_ denotes the number of individuals who transition from the infected class *I* to the reported class *I*_*q*_ per day; it is also directly related to the daily number of COVID-19 tests carried out during the outbreak.

**Figure 1:**
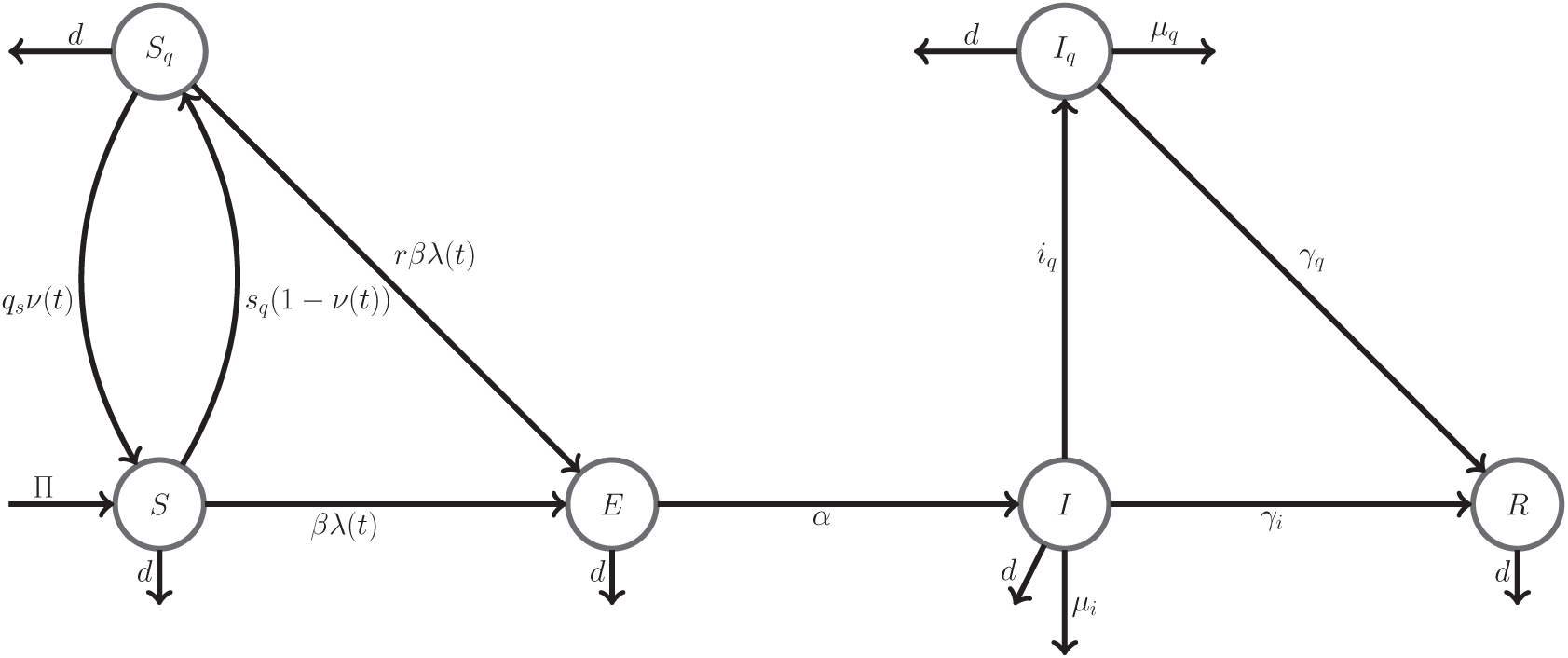
Flow diagram illustrating the disease transitions among the compartments

Susceptible individuals make the transition to the *S*_*q*_(*t*) compartment with a rate of 

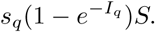

Note that the main indicator of quarantine is the number of reported cases *I*_*q*_. When the number of reported cases increases in a state or country, then the quarantine is imposed or naturally taken as an option. Thus, when the number of reported cases increases, then percentage or amount of people quarantined will increase. If the number of reported cases falls to zero, the transition rate from *S* to *S*_*q*_ is zero and from *S*_*q*_ to *S* is *q*_*s*_. The individuals in *S* and *S*_*q*_ compartments transition to compartment *E* (exposed) with a force of infection given by 

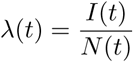

and disease transmission rate *β*. Note that, since the individuals in *S*_*q*_ transition to *E* compartment less frequently, a reduction factor *r* is taken into count in the model. After an incubation period of 1*/α*, the individuals in *E* compartment transition to *I* compartment (infected) with rate *α*. The individuals in *I* compartment will either transition to *R* compartment (recovered) with a rate of *γ*_*i*_ or *I*_*q*_ compartment with a rate of *i*_*q*_, or die due to the disease with a rate of *µ*_*i*_. The individuals in *I*_*q*_ (reported (infected) individuals, who are hospitalized or quarantined) compartment either transition to *R* compartment with a rate of *γ*_*q*_ or die due to disease with a rate of *µ*_*q*_. The following (ODEs) system represents dynamical behavior of the system. 

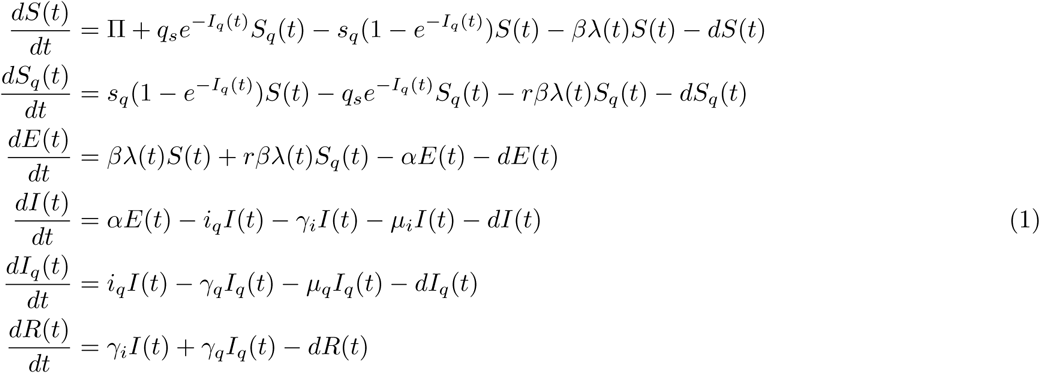

with *S*(0) *>* 0, *S*_*q*_(0) *≥* 0, *E*(0) *≥* 0, *I*(0) *≥* 0, *I*_*q*_(0) *≥* 0, *R*(0) *≥* 0 and *N* (*t*) = *S*(*t*) + *S*_*q*_(*t*) + *E*(*t*) + *I*(*t*) + *I*_*q*_(*t*) + *R*(*t*) The left hand side of the system (1) represents the rate of change per day. In the system (1), we have 

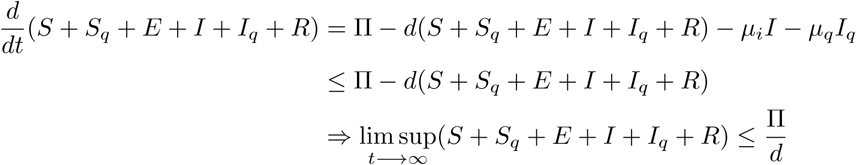

Hence, the feasible region of the system (1) is given by 

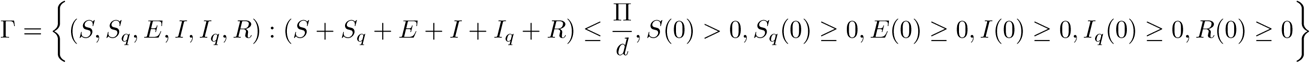

This implies all the compartments stay non-negative. The parameter values used in the model are given in Table 1 with their description and units.

**Table 1:** Parameter descriptions and values

| Parameter | Description | Value | Unit | Source |
| --- | --- | --- | --- | --- |
| $\mathcal{R}_0$ | The basic reproductive number | 5.49 | - | Estimated |
| $\beta$ | Disease transmission rate | 5.24 | $\frac{1}{\text{day}}$ | Estimated |
| $q_s$ | Rate of release from quarantine | 0.5 | $\frac{1}{\text{day}}$ | Estimated |
| $s_q$ | Self-quarantine rate of susceptible individuals | 0.096 | $\frac{1}{\text{day}}$ | Estimated |
| $\alpha$ | Rate of exposed individuals becoming infected | 0.2381 | $\frac{1}{\text{day}}$ | (Nishiura, Linton, & Akhmetzhanov, 2020; Guan et al., 2020; Sanche et al., 2020) |
| $\gamma_i$ | Recovery rate due to natural immune response | 0.111 | - | Estimated |
| $\gamma_q$ | Recovery rate due to hospitalization | 0.107 | $\frac{1}{\text{day}}$ | Estimated |
| $\mu_i$ | Death rate of disease in infectious compartment | 0.0096 | $\frac{1}{\text{day}}$ | Estimated |
| $\mu_q$ | Death rate due to disease in reported infectious compartment | 0.004 | $\frac{1}{\text{day}}$ | Estimated |
| $i_q$ | Rate of reported (infected) individuals | 0.832 | $\frac{1}{\text{day}}$ | Estimated |
| $r$ | Reduction rate in transmission due to self quarantine | 0.011 | - | Estimated |
| $\Pi$ | Recruitment rate | 2134 | $\frac{\text{Individual}}{\text{day}}$ | (World Health Organization, 2020a) |
| $d$ | Natural death rate | 0.000023 | $\frac{1}{\text{day}}$ | (World Health Organization, 2020a) |
| $E(0)$ | Initial number of exposed Individuals | 142 | Individual | Estimated |
| $I(0)$ | Initial number of infected Individuals | 69 | Individual | Estimated |

## 3 Diseases free equilibrium and stability analysis

One of major concepts in an outbreak is disease free equilibrium (DFE), where the entire population is susceptible (Keeling & Rohani, 2008; Diekmann, Heesterbeek, & Roberts, 2010). For the system (1), the DFE can be denoted 

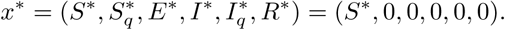

To able to get the DFE for the system (1), we set the right hand side of the system (1) to zero and substitute the DFE into the system. Hence, the DFE is found as 

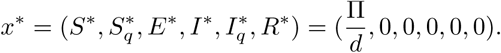

We then analyze whether the DFE is stable or not. Next-generation matrix (NGM) is used (Van den Driessche & Watmough, 2002; Diekmann et al., 2010; Van den Driessche & Watmough, 2008) to determine the stability of DFE. We rewrite our system (1) as:

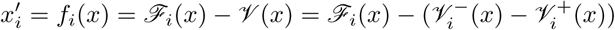

where 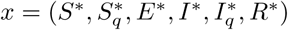and *i* = 1, …, 6 and

- *ℱ*_*i*_(*x*):= Rate of appearance of new infections in compartment *i*
- 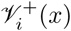:=Rate of transfer of individuals into compartment *i* by all other means
- 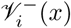= Rate of transfer of individuals out of compartment *i* by all other means. Hence, *ℱ* and *𝒱* are calculated for the system of (1):

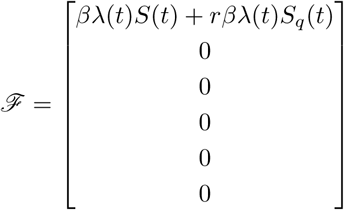

And 

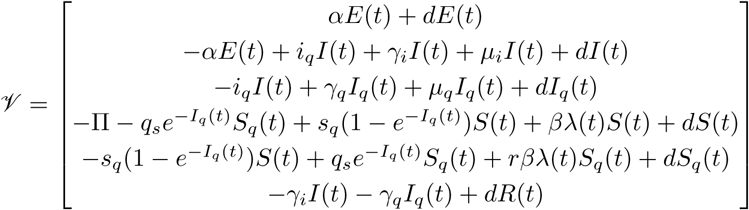

Notice that individuals which transition to *E* compartment are the only newly infected cases. Therefore, the Jacobian at the DFE for the infected classes (first three components, *x*) are 

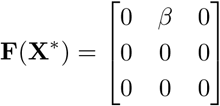

and 

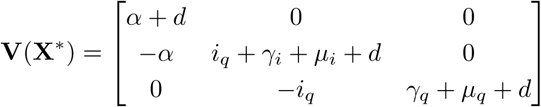

Then, we compute the next generation matrix (NGM) as 

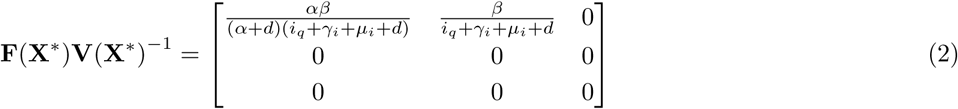

The spectral radius of the NGM is the basic reproduction number, *ℛ*_0_ defined as the average number of secondary cases arising from an average primary infected case in an entirely susceptible population. The DFE is locally stable if *ℛ*_0_ *<* 1 (Van den Driessche & Watmough, 2002; Diekmann et al., 2010). The spectral radius of the NGM given in (2) 

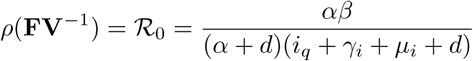

Since we do not consider the individuals in *S*_*q*_ compartment as a part of the DFE, we do not see any effect of quarantine on *ℛ*_0_. However, *s*_*q*_ indirectly changes the other parameter values such as *β, i*_*q*_, therefore, *ℛ*_0_ value changes with *s*_*q*_ indirectly. Note that *α, β* are positively correlated with *ℛ*_0_ and *i*_*q*_, *γ*_*i*_, *µ*_*i*_, *d* are negatively correlated with *ℛ*_0_ for the system (1). Note that we might control the disease with increasing quarantine rate of infected individual *i*_*q*_. Thus, if 

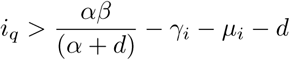

then the DFE is locally stable and the disease dies out when sufficiently close to DFE. Biologically, if the infected individuals can be detected in a sufficiently short time another word, the number of test to detect the number of cases increase, then the disease can be controlled.

## 4 Parameter estimation

In this part, we estimate the parameters in the system (1), so we fit our model with the daily reported cumulative number of cases and deaths, which are provided by (World Health Organization, 2020b) and (Chinese physicians, 2020). We use the Ordinary Least Squares (OLS) method and minimize the sum of the squares of differences between the daily reported data and those predicted by our model. The goodness of the fit is measured by computing the associated relative error of the fit using the formula 

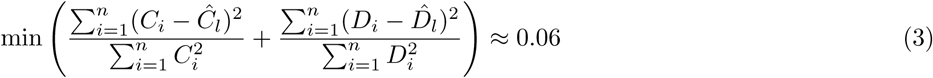

where *C*_*i*_ and *Ĉ*_*l*_ are exact and estimated cumulative(infected) cases, and *D*_*i*_ and 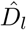 are exact and estimated cumulative deaths. To estimate the number of COVID-19 deaths, we sum the number of deaths coming from the infected class *I* and the reported (infected) class *I*_*q*_. Note that the natural deaths in the infected class *I* and the reported (infected) class *I*_*q*_ are also included in the total number of deaths. We used an ode45 solver with fmincon from the Optimization Toolbox of MATLAB. By using the initial conditions: *S*(0) = 59, 000, 000, *S*_*q*_(0) = 0, *I*_*q*_(0) = 258, and *R*(0) = 0, we estimate all the parameters of the model together with estimating the initial number of exposed *E* and infected *I*, except the natural death rate *d*, recruitment rate II, and incubation period *α*. We used 4.2 days for the average incubation period that is provided by (Nishiura et al., 2020; Guan et al., 2020; Sanche et al., 2020). The natural death and recruitment rate are provided by (World Health Organization, 2020a).

The simulation results obtained for the cumulative number of (infected) cases *C* and cumulative deaths *D* by fitting the model with the data from January 20, 2020 to March 23, 2020 are depicted in Figure 2. These figures show a reasonably good fit with the total relative error 0.06 (6%). Most of the error comes from the fit of cumulative cases, especially around February 12, 2020. In February, China began to report clinically diagnosed cases in addition to laboratory-confirmed cases, and on February 12, 2020, 13,332 clinically (rather than laboratory) cases reported even though they were diagnosed in the preceding days and weeks. Due to the very small number of cases reported after March 23, 2020, we chose to fit the model using only data from before this date.

**Figure 2:**
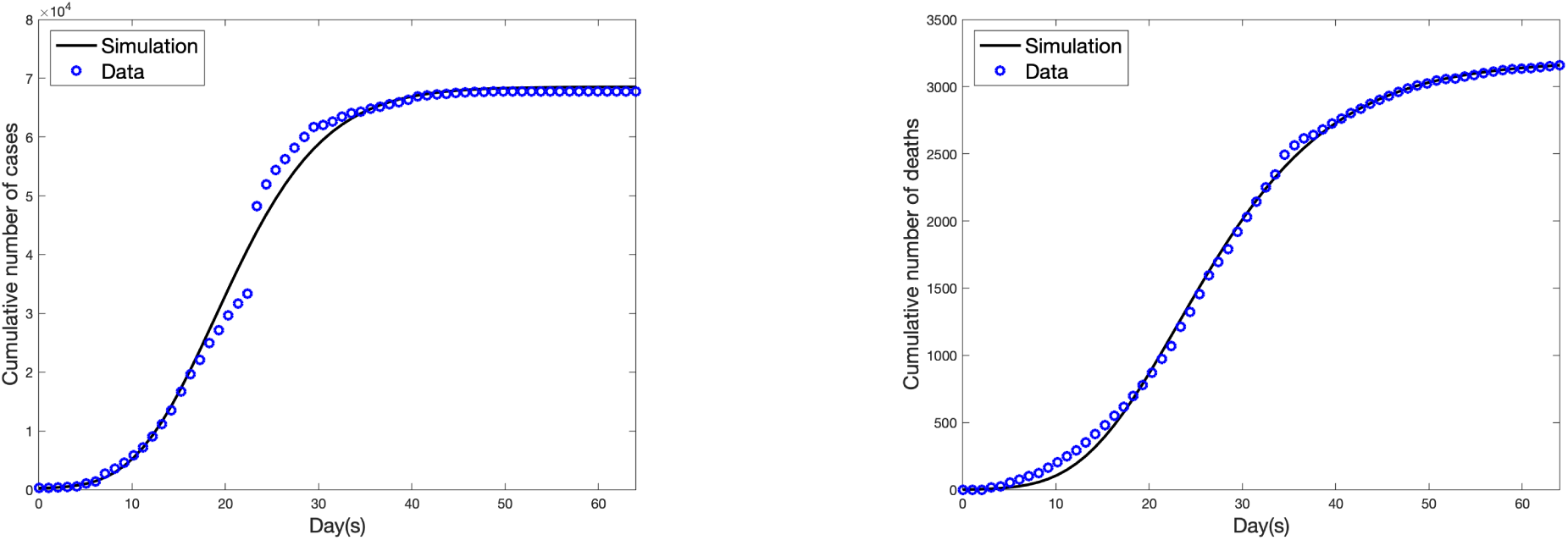
Fitting the model to the data from January 20, 2020 to March 23, 2020 in Hubei: the cumulative number of (infected) cases and deaths (Parameter values used are as given in Table 1).

## 5 Simulations

In this section, we discuss 15-day forecasting of the outbreak, the effect of quarantine, and the effect of testing in Hubei. We also conduct a sensitivity analysis to see which parameters play important role in the dynamics of the outbreak.

### 5.1 Effect of quarantine (Self-isolation)

When we look at the change in the quarantine class, nearly the entire province of Hubei was quarantined by February 15, 2020 (See Figure 3, February 15 corresponds the day 25 in the figure). The percentage of the population transitioning from the susceptible class to the quarantine class attains its maximum level between January 27, 2020 and February 10, 2020, and by February 15, almost all of the population were in quarantine. This result makes sense since the state government imposed a quarantine in the state on January 23, 2020, initially recommending quarantine and finally forcing the people into quarantine to guarantee social distancing. This action seems to have worked to great effect, reducing the contact rate by about 98.9% (See Table 1 for the parameter, *r*, the reduction rate due to the quarantine).

**Figure 3:**
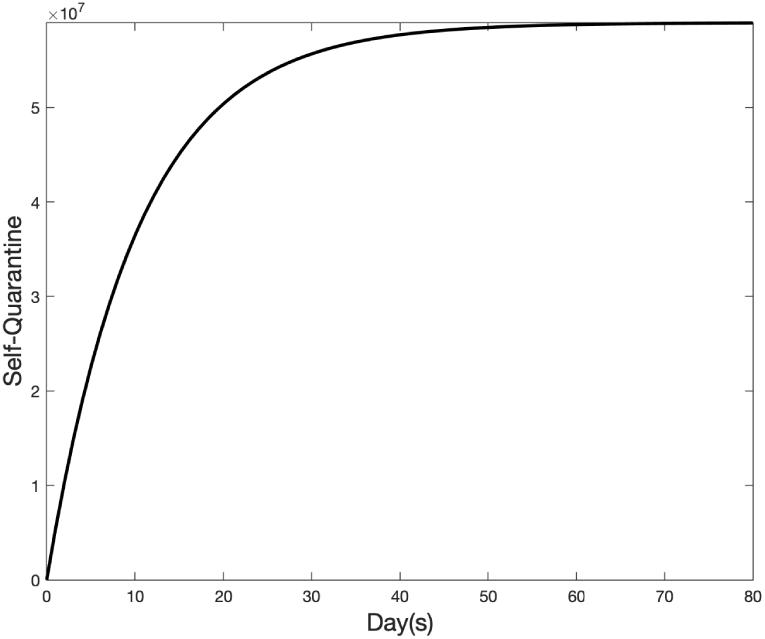
Transitions from the susceptible class to the quarantine class during the outbreak in Hubei

When we reduce the quarantine rate, *s*_*q*_ from 0.096 to 0.0864 (10% reduction) and do not change the remaining parameters, the number of cases and deaths would be about 141090 and 6562, respectively. Similarly, when we increase the quarantine rate, *s*_*q*_ from 0.096 to 0.1056 (10% increase), the number of cases and deaths would be about 39334 and 1829, respectively. Thus, any change in the quarantine rate makes very significant change in total number of cases and deaths. Furthermore, see the sensitivity analysis section below, the quarantine rate is a significant parameter in the dynamics of the outbreak as well as its efficiency, which is explained by the parameter, r is also significant in the dynamic of the outbreak.

### 5.2 15-day Forecasting

We used parameters in Table 1 for 15-day forecasting. Figure 4 shows the estimated number of infected cases for 80 days. The plot on the left depicts the estimated number of exposed and the right plot depicts the estimated number of reported (infected) cases *I*_*q*_. As it can be seen, the number of individuals in each of these classes tends to zero, which implies that the outbreak is almost over, and so new cases may not be recorded in Hubei. As it can be seen from the change in infected class, the outbreak reaches its peak about February 9, 2020. The infected class *I* also shows how many people were out with no symptoms or mild symptoms during the outbreak.

**Figure 4:**
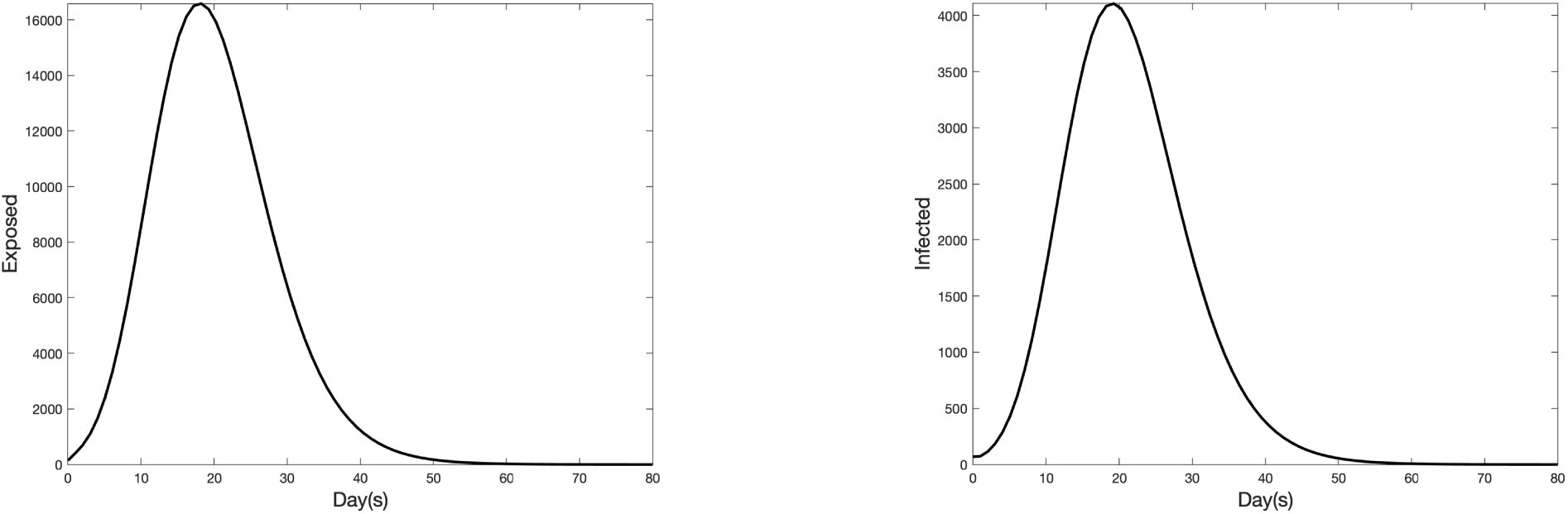
The plot on the left depicts the number of exposed cases and the plot on the right depicts the number of infected cases with initial conditions *S*(0) = 59, 000, 000, *S*_*q*_(0) = 0, *E*(0) = 142, *I*(0) = 69, *I*_*q*_(0) = 258, *R*(0) = 0 for 80 days

In Figure 5, the plot on the left shows the estimated number of cumulative reported (infected) cases and the right plot shows the estimated number of cumulative deaths. As of 30 March 2020, there were no reported cases in Hubei in the past week and the total number of cases and total number of deaths were 67801 and 3187, respectively. Our model (1), predicts the number of cases and deaths with high accuracy with 6 percent relative error. We estimated the fatality rate of the outbreak in Hubei as approximately 4.8% with the estimated number of cases, about 67994 and deaths, about 3254.

**Figure 5:**
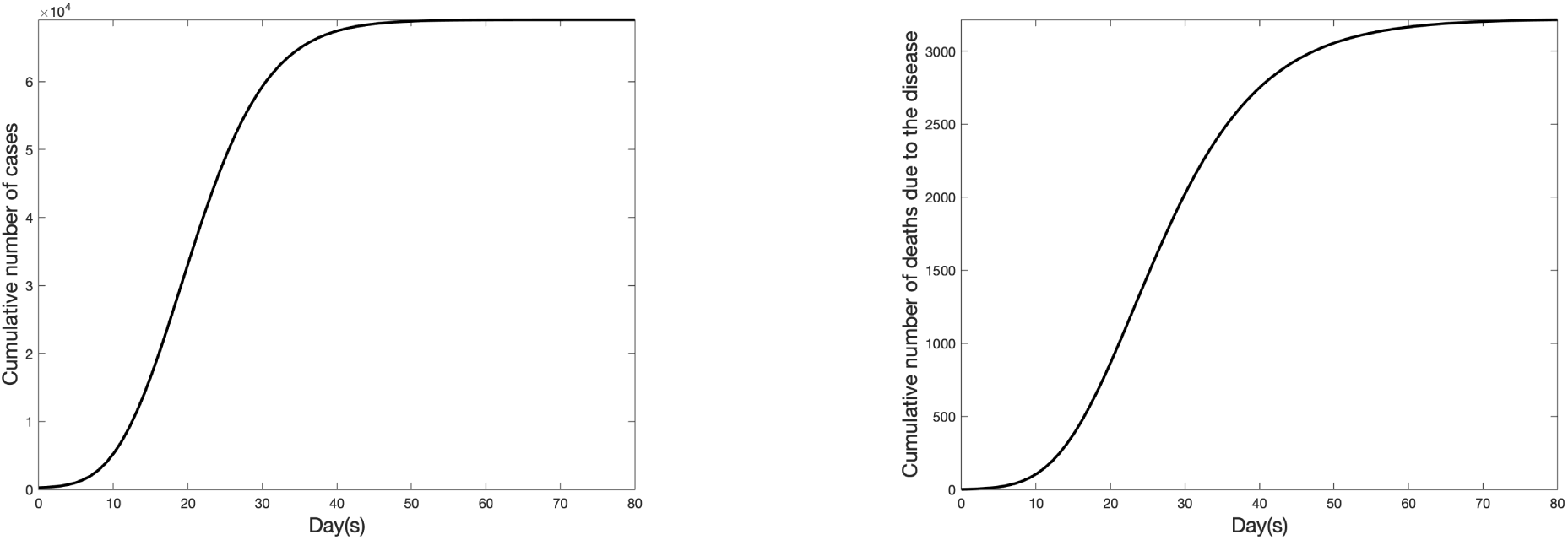
The plot on the left depicts the simulated cumulative number of infected cases due to the outbreak and the plot on the right depicts the simulated cumulative number of deaths in Hubei

### 5.3 Sensitivity analysis

Several parameters play important roles in the model (1). These parameters were estimated with existing data as of (Coronavirus COVID-19 Global Cases by Johns Hopkins CSSE, 2020). In order to determine the set of parameters that are statistically significant regarding the number of cumulative infected cases, we conduct a sensitivity analysis of the model. We utilized a Latin Hypercube Sampling (LHS) and the Partial Rank Correlation Coefficients (PRCC) method (Marino, Hogue, Ray, & Kirschner, 2008). We use a range given in Table 2 to sample parameters from a uniform distribution, then use these samples as input variables when we run the system (1) with initial conditions *S*(0) = 1000, *S*_*q*_(0) = 0, *E*(0) = 10, *I*(0) = 3, *I*_*q*_(0) = 0, *R*(0) = 0 for 90 days. The number of cumulative infected cases is the output variables in sensitivity analysis. Table 2 shows PRCC values, p-values and the range for each corresponding parameters.

**Table 2:** Results of sensitivity analysis with partial rank correlation coefficient (PRCC) and p-value with the range of parameters that we made the sensitivity analysis

| Parameter | PRCC | p-value | Range |
| --- | --- | --- | --- |
| $\beta$ | 0.89 | 1.2e-104 | [1, 10] |
| $q_s$ | -0.084 | 0.15 | [0.1, 0.9] |
| $s_q$ | -0.45 | 3.4e-16 | [0.01, 0.2] |
| $\alpha$ | 0.23 | 6.9e-05 | [0.05, 0.5] |
| $\gamma_i$ | -0.25 | 1.3e-05 | [0.01, 0.2] |
| $\gamma_q$ | -0.075 | 0.2 | [0.01, 0.4] |
| $\mu_i$ | -0.052 | 0.37 | [0.001, 0.2] |
| $\mu_q$ | 0.12 | 0.032 | [0.001, 0.2] |
| $i_q$ | -0.81 | 3.1e-70 | [0.5, 4] |
| $r$ | 0.69 | 8.3e-44 | [0.001, 0.9] |
| $\Pi$ | -0.05 | 0.39 | [0.01, 0.1] |
| $d$ | 0.067 | 0.25 | [0.000001, 0.0001] |

The sensitivity analysis indicates that *β, s*_*q*_, *i*_*q*_, and *r* are statistically more significant parameters depending on the high PRCC values in the dynamics of the outbreak. Therefore, it is of interest to study how the number of cumulative infected cases changes when *s*_*q*_, *i*_*q*_, *r*, and *β* are varied and other parameters are held the same as in Table 1 and the initial condition same as before. Figure 6 shows the results of these experiments, how the number of cumulative cases changes for different values of *s*_*q*_, *i*_*q*_, *r*, and *β*.

**Figure 6:**
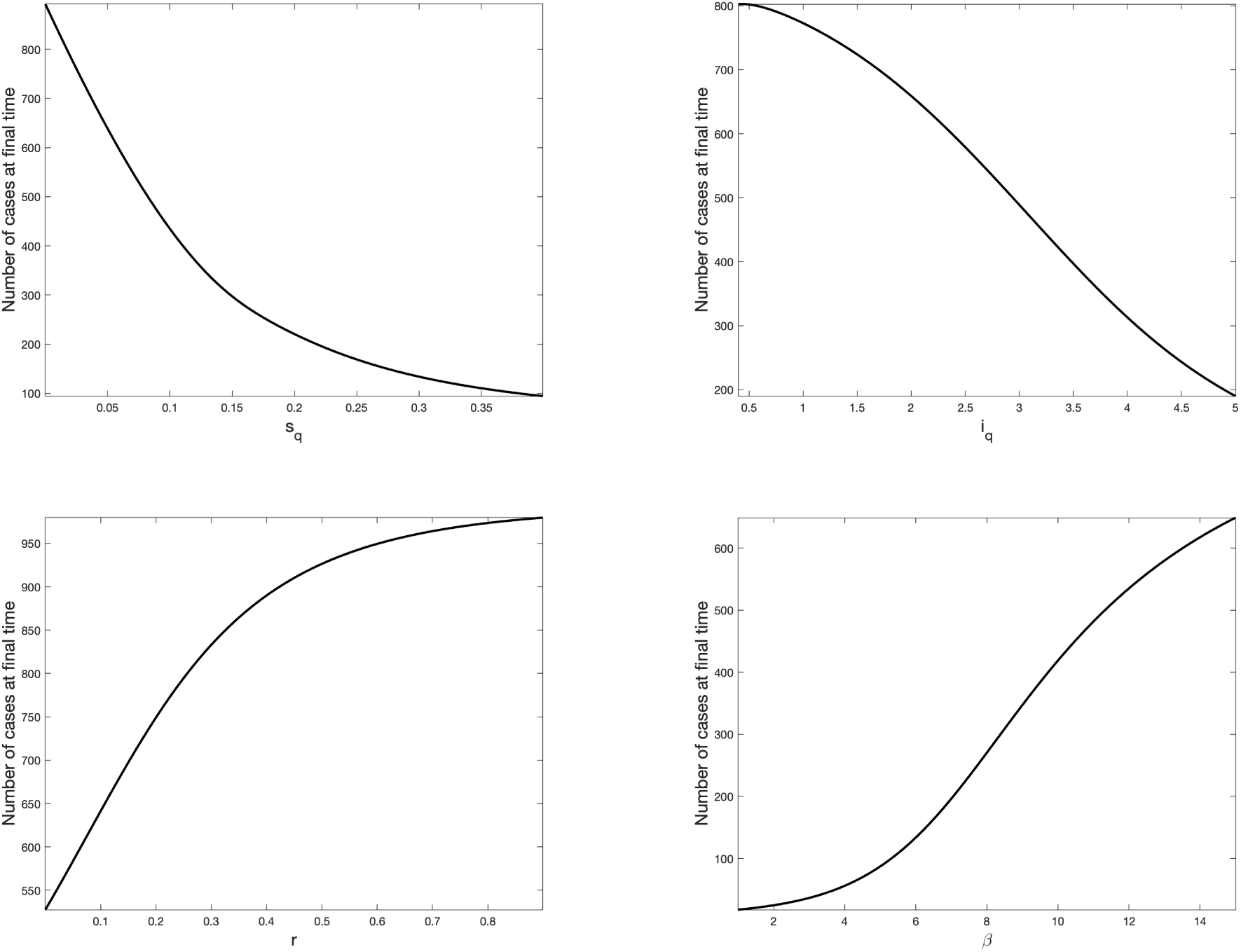
Number of cases at final time with respect to each parameters

Figure 6 shows that an increase in the quarantine rate, *s*_*q*_, sharply decreases the number of cumulative cases, in range [0.005, 0.2] and the quarantine rate of infected individuals, *i*_*q*_, decreases the number of new cases mostly in range [1.5, 4.5]. Increasing the reduction rate *r*, due to the quarantine, increases the number of cumulative cases mostly in range [0.1, 0.6] and the disease transmission rate, *β* increases the number of cases mostly in range [4, 10].

It is also important to analyze how *ℛ*_0_ value varies with *β* and *i*_*q*_. Thus, we vary *β* in the range [1, 10] and *i*_*q*_ in the range [0.1, 4] while keeping all other parameters the same in Table 1. Figure 7 shows the boxplot of *β* and *i*_*q*_. We observe *i*_*q*_ affects *ℛ*_0_ in a wider range compare to *β*. Thus, the range of *ℛ*_0_ will change roughly between 3 and 8. In addition, the value of *ℛ*_0_ drops below 1 when *i*_*q*_ is above 5.1.

**Figure 7:**
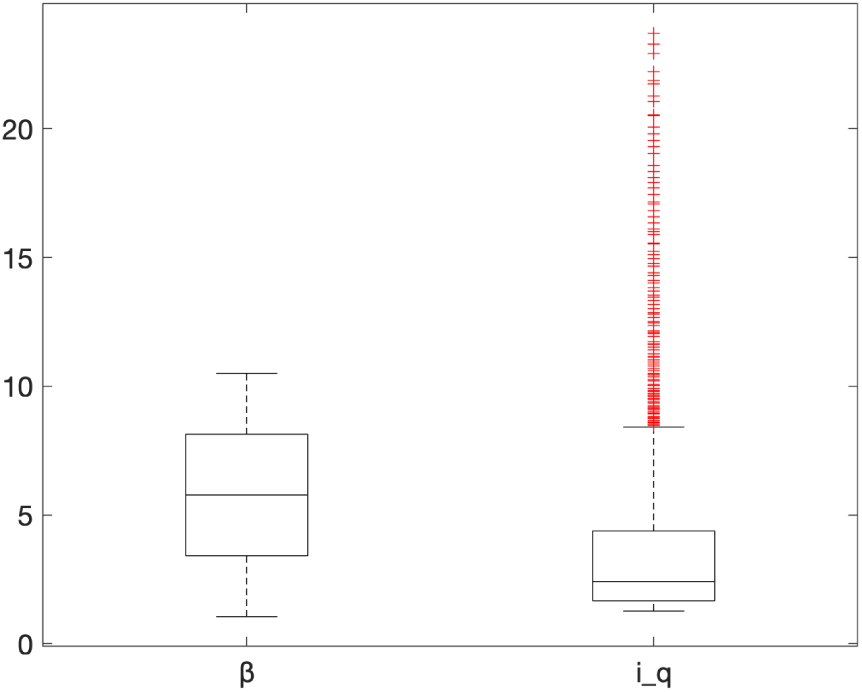
Change in *ℛ*_0_ when we vary *β* with the rage [1,10], or *i*_*q*_ with the range [0.1, 4].

The rate of reported (infected) cases, *i*_*q*_ is related to the number of tests given during the outbreak to identify the infected people. Thus, increasing the number of tests will increase the rate of case reporting *i*_*q*_. This will reduce the number of cases (see Figure 6) and, consequently, the number of deaths due to the outbreak. When we increase the rate of reported (infected) cases *i*_*q*_ by about 10%, the number of cases and number of deaths are estimated to be 36040 and 1639, respectively. Decreasing the rate about 10%, the number of cases and number of deaths are estimated to be 14084 and 6724, respectively.

## 6 Forecasting the peak of the outbreak, number of cases/deaths in Turkey

In the part, we fit the model (1) with available COVID-19 data from Turkey ((Ministry of Health (Turkey), 2020)). We fit the model (1) with Turkish data from March 10, 2020 to April 10, 2020, and get about 5.9% relative error in the fit by using the equation (3). We estimate the four parameters *i*_*q*_, *s*_*q*_, *β*, and *r*, which are not only the most significant parameters in the dynamics of outbreak, but also are specific to each country since they are related to the number of COVID-19 tests administered *i*_*q*_, the number of individuals in quarantine *s*_*q*_, the contact rate of individuals *β*, and the efficiency of quarantine *r* in each country. Therefore, by using the initial conditions: *S*(0) = 83, 000, 000, *S*_*q*_(0) = 0, *I*_*q*_(0) = 1, and *R*(0) = 0, we estimate these four parameters together with the initial number of exposed and infected individuals. We do not estimate the rest of the parameters, employing the parameters in Table 1. Therefore, our results in this section will depend on observed dynamics of the outbreak in Hubei as well as the available Turkish data (((Ministry of Health (Turkey), 2020)). Note that the quarantine rate and the rate of reported cases (which, we stress, is related to number of COVID-19 tests) can be increased, and the increase still may have significant effect toward the reduction of the number of cases (See Figure 6, sensitivity analysis), but increasing the reduction rate *r* does not make very significant changes by way of the number of cases in Turkey since it is very close to its maximum level (See Figure 6, sensitivity analysis part). Thus, we will vary only the quarantine rate *s*_*q*_ and the rate of reported cases *i*_*q*_ in forecasting the peak of the outbreak and the number of cases/deaths in Turkey.

The rate of reported cases is about 1.8; this rate is larger than what we observed in Hubei. This implies that in terms of numbers of COVID-19 tests conducted per day, Turkey is now doing a better job than Hubei, China at a comparable time in Hubei’s outbreak. The efficiency of quarantine also seems to be very good in Turkey, given the approximately 85% reduction in the contact rate of COVID-19 obtained by our parameter estimation. On the other hand, the quarantine rate is about 0.088, which is small when compared with the quarantine rate in Hubei (the rate was 0.096 in Hubei). In Hubei, the population transitioned to quarantine class very quickly (almost in two weeks), but in Turkey the movement to quarantine has been very slow in comparison (see Figure 8), suggesting why the contact rate is higher in Turkey when we compare to the contact rate in Hubei (See Table 1 and 3 for these rates).

**Table 3:** Parameter descriptions and values of the model (1) with Turkish data.

| Parameter | Description | Value | Unit | Source |
| --- | --- | --- | --- | --- |
| $\mathcal{R}_0$ | The basic reproductive number | 5.2 | - | Estimated |
| $\beta$ | Disease transmission rate | 9.98 | $\frac{1}{\text{day}}$ | Estimated |
| $s_q$ | Self-quarantine rate of susceptible individuals | 0.088 | $\frac{1}{\text{day}}$ | Estimated |
| $i_q$ | Rate of reported (infected) individuals | 1.8 | $\frac{1}{\text{day}}$ | Estimated |
| $r$ | Reduction rate in transmission due to quarantine | 0.15 | - | Estimated |
| $\Pi$ | Recruitment rate | 3827 | $\frac{\text{Individual}}{\text{day}}$ | (World Health Organization, 2020a) |
| $d$ | Natural death rate | 0.000016 | $\frac{1}{\text{day}}$ | (World Health Organization, 2020a) |
| $E(0)$ | Initial number of exposed Individuals | 6 | Individual | Estimated |
| $I(0)$ | Initial number of infected Individuals | 2 | Individual | Estimated |

**Figure 8:**
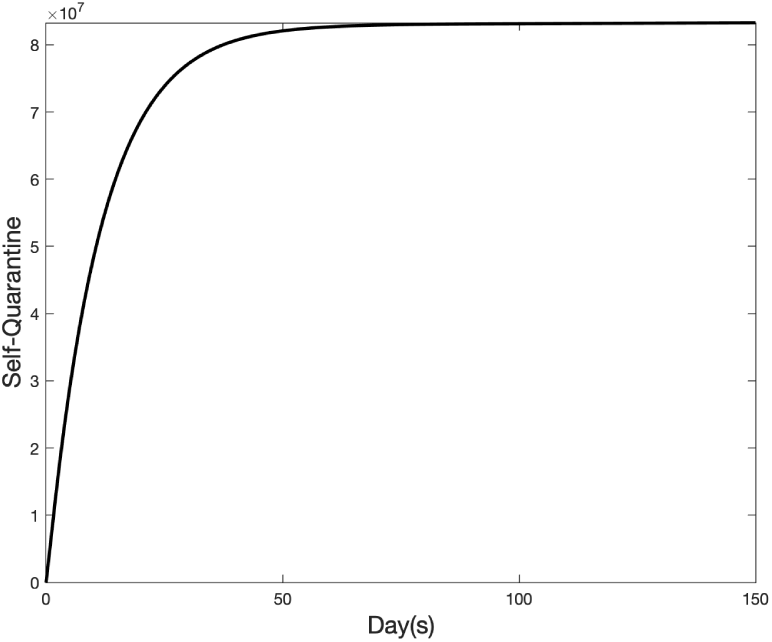
Transitions from the susceptible class to the quarantine class during the outbreak in Turkey after the first cases reported on March 10, 2020

It is still possible to increase the quarantine rate (the rate, per day, of transition to quarantine class) and the number of COVID-19 tests given each day in Turkey to make a reduction in the number of cases and deaths (See Figures 9 and 10). In Figures 9 and 10, the red curves are obtained using base parameters from Table 1 and 3, and the other curves obtained by varying the quarantine rate *s*_*q*_ and the rate of reported cases *i*_*q*_. When we use the base parameter values which are obtained from our fitting, Turkey then will have about 203,700 cases and 8,269 deaths. If Turkey can increase the number of individuals in quarantine and the number of daily COVID-19 tests, then, depending on the magnitude of the increases, the number of cases and deaths can decrease significantly (see Figures 9 and 10). When we look at trajectories of cumulative cases and deaths in Figures 9 and 10, in the worst-case scenario (the black curves) of the study, Turkey will have about 281,500 cases and 11,430 deaths. These projections decrease to 148,100 cases and 6,005 deaths if Turkey can increase the number of individuals in quarantine and the number of COVID-19 tests.

**Figure 9:**
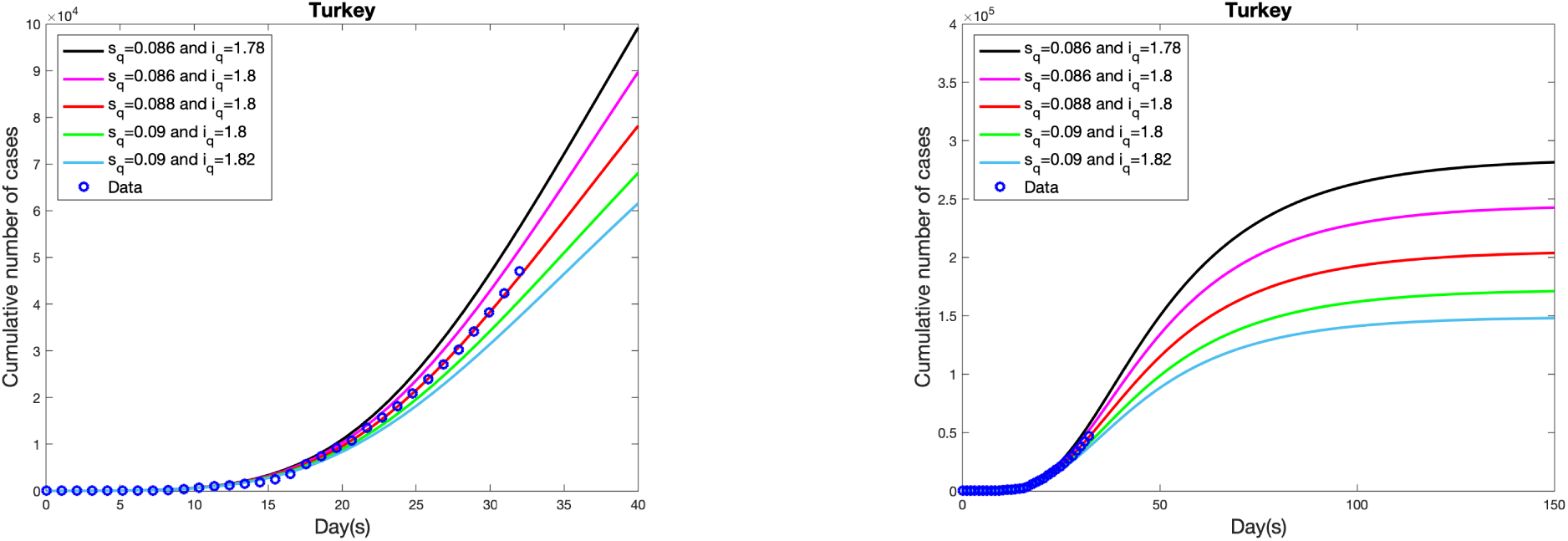
Cumulative number of (infected) cases depending on different quarantine rate *s*_*q*_ and rate of reported cases *i*_*q*_. Left graph shows the cumulative number of cases between day 1 to day 40, and right plot shows the cumulative number of cases between day 1 to day 150 in the outbreak in Turkey.

**Figure 10:**
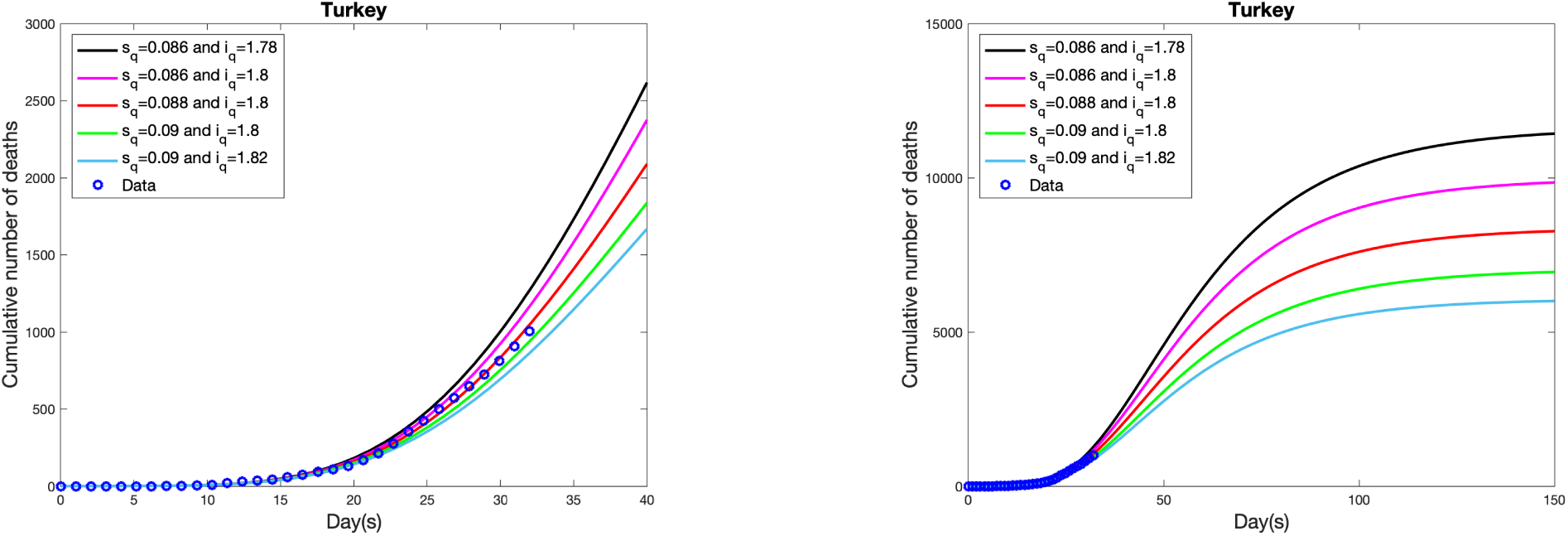
Cumulative number of deaths depending on different quarantine rate *s*_*q*_ and rate of reported cases *i*_*q*_. Left graph shows the cumulative number of deaths between day 1 to day 40, and right plot shows the cumulative number of deaths between day 1 to day 150 in the outbreak in Turkey.

The peak of the outbreak in Turkey is also very sensitive to the quarantine rate *s*_*q*_ and the rate of reported cases *i*_*q*_. Depending on the change in quarantine rate and the rate of reported cases *i*_*q*_, the peak of outbreak in Turkey can be seen between the day 42 (April 20,2020) and day 48 (April 26, 2020), and the outbreak will almost die out by the day 150 (at the end of July 2020, see Figure 11).

**Figure 11:**
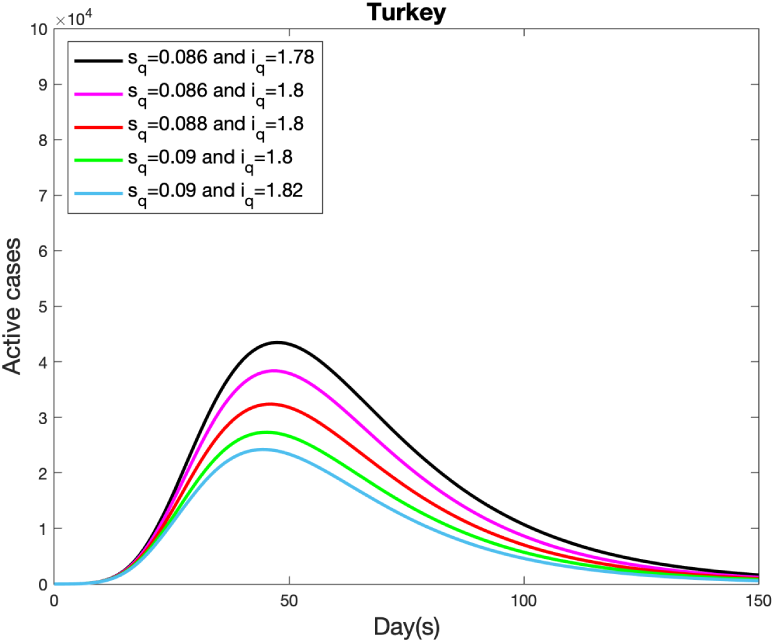
Projected (simulated) peak of outbreak in Turkey depending on different quarantine rate *s*_*q*_ and rate of reported cases, *i*_*q*_

## 7 Conclusion and Discussion

Our analysis suggests that quarantine greatly reduced the number of cases and deaths seen in Hubei’s COVID-19 outbreak. In addition, while quarantine does not appear in the representation of *ℛ*_0_, it still indirectly reduces *ℛ*_0_. We also saw that the dynamics of the outbreak is very sensitive to the quarantine rate *s*_*q*_ and contact rate *β*, as indicated by our sensitivity analysis. The basic reproductive number is estimated as 5.49 and the study shows that any change in *β* or *i*_*q*_ directly affects the basic reproductive number.

The quarantine decidedly reduces the number of cases and deaths. Increasing (or decreasing) the speed of movement from the susceptible class to the quarantine class by about 10% would double (or half) the number of cases and deaths due to the outbreak (This speed of movement is controlled by the rate *s*_*q*_). Of course, the efficiency of the quarantine is also very important. In our model, the efficiency of the quarantine measured by the reduction rate, *r*. The reduction rate shows how much reduction is effected in the contact of COVID-19 thanks to the quarantine. Based on our sensitivity analysis, this parameter is very important (see Figure 6). Our model shows that the quarantine in Hubei was almost perfect since it caused about 98.9 percent reduction in the contact rate of COVID-19.

Another important parameter that plays a crucial role in the dynamics of the outbreak is the rate of reported (infected) cases *i*_*q*_ which is directly related to the number of tests given to detect infected individuals. Similar to the quarantine rate *s*_*q*_, the rate of reported (infected) cases *i*_*q*_ could double (half) when we have 10% reduction (or increase) in the rate.

As of 30 March 2020, there were no reported cases in Hubei in the past week and the total number of cases and total number of deaths were 67801 and 3187, respectively. Based on our 15-days forecasting, the number of cases in Hubei was projected to be about 67994 and the number of deaths was projected to be about 3254. Thus, we estimate the fatality rate of the outbreak to be about 4.8% in Hubei. Our model gives about 6% relative error and we are confident that using the model will be helpful for forecasting local outbreaks of the pandemic in other regions.

From existing COVID-19 data from Turkey and the dynamics of our model understood from the Hubei analysis, the outbreak in Turkey is expected to reach its peak between April 20 and April 26 depending on the number of individuals (amount of people) in quarantine and the number of COVID-19 tests carried out each day in Turkey. The daily number of tests given in Turkey is large when we compare to the rates of reported cases in Hubei. As we showed in the sensitivity analysis, increasing the number of COVID-19 tests and the number of individuals in quarantine will significantly reduce the number of cases (and deaths). Based on our forecasting, the number of cases will be about 203,700 with the range 148,100 and 281,500, and the number of deaths will be about 8,269 with the range 6,005 and 11,430 depending on quarantine rate, *s*_*q*_ and the rate of reported cases, *i*_*q*_ in Turkey. Thus, in any cases that are given in Figure 9 and 10, the fatality rate of COVID-19 will be about 4.1% in Turkey.

Based on the results above, our main concern in Turkey is the proportion of people in quarantine. The quarantine rate is 0.088 in Turkey, a low number when we compare with the quarantine rate in Hubei, China (it was 0.096 in Hubei). If Turkey can increase the number of individuals in quarantine, then the number of cases and deaths may decrease significantly (See Figure 9 and 10 to see the effect of quarantine in COVID-19 cases and deaths). Small changes in quarantine rate make significant changes in the total number of cases and deaths in Turkey.

The efficiency of the quarantine in Turkey is about 85 percent, meaning that it causes 85 percent reduction in the contact rate of COVID-19. Thus, the quarantine rate, *s*_*q*_ and its efficiency is very important to be able to contain the outbreak (See Table 3 for reduction rate, *r* and Figure 6 for the effect of reduction rate in the total number of cases).

As of April 10, 2020, the number of COVID-19 tests given each day in Turkey had increased to 30,000 ((Ministry of Health (Turkey), 2020)). If the number is increased further, then it also will decrease the total number of cases and deaths in Turkey (See Figure 9 and 10). Indeed, it is expected that the number of cases may drop below to 203,700 and stay between 148,100 and 203,700. Similarly, this could cause the number of deaths in Turkey to decrease below 8,269 and stay between 6,005 and 8,269.

## Data Availability

World Health Organization
Coronavirus COVID-19 Global Cases by Johns Hopkins CSSE. (2020)
Chinese physicians. (2020). ncov.dxy.cn.
Ministry of Health (Turkey). (2020). Coronavirus disease 2019 (covid-19) daily data.
World Health Organization. (2020b). novel coronavirus (covid-2019) situation reports.

https://www.who.int/

https://www.who.int/emergencies/diseases/novel-coronavirus-2019/situation-reports

https://ncov.dxy.cn/ncovh5/view/pneumonia?scene=2&clicktime=1579582238&enterid=1579582238&from=singlemessage&isappinstalled=0https://ncov.dxy.cn/ncovh5/view/pneumonia?scene=2&clicktime=1579582238&enterid=1579582238&from=singlemessage&isappinstalled=0

https://gisanddata.maps.arcgis.com/apps/opsdashboard/index.html#/bda7594740fd40299423467b48e9ecf6

https://covid19.saglik.gov.tr

## Authors’ Contributions

All authors contributed equally to this work.

## Acknowledgements

The authors would like to acknowledge the generous support of the Turkish Ministry of National Education in the study.

